# Convalescent plasma for patients with severe COVID-19: a matched cohort study

**DOI:** 10.1101/2020.08.18.20177402

**Authors:** Ralph Rogers, Fadi Shehadeh, Evangelia Mylona, Josiah Rich, Marguerite Neill, Francine Touzard-Romo, Sara Geffert, Jerome Larkin, Jeffrey A. Bailey, Shaolei Lu, Joseph Sweeney, Eleftherios Mylonakis

## Abstract

**Background:** The efficacy of convalescent plasma (CP) for the treatment of COVID-19 remains unclear.

**Methods:** A matched cohort analysis of hospitalized patients with severe COVID-19. The impact of CP treatment on all cause in-hospital mortality was evaluated using univariate and multivariate Cox proportional-hazards models, and the impact of CP treatment on the time to hospital discharge was assessed using a stratified log-rank analysis.

**Results:** 64 patients who received CP a median of 7 days after symptom onset were compared to a matched control group of 177 patients. Overall in-hospital mortality was 14.9%. There was no significant difference in the risk of in-hospital mortality between the two groups (adjusted hazard ratio [aHR] 0.93, 95% confidence interval [CI] 0.39 – 2.20). There was also no significant difference in the overall rate of hospital discharge (rate ratio [RR] 1.28, 95% CI 0.91 – 1.81), but a subgroup analysis of patients 65-years-old or greater who received CP demonstrated a significantly increased hospital discharge rate among these patients (RR 1.86, 95% CI 1.03 – 3.36). There was a greater than expected frequency of transfusion reactions in the CP group (2.8% reaction rate observed per unit transfused).

**Conclusions:** The use of CP in this study was a safe treatment for COVID-19. There was no overall significant reduction of in-hospital mortality or increased rate of hospital discharge associated with the use of CP in this study, although there was a signal for improved outcomes among the elderly. Further adequately powered randomized studies should target this subgroup when assessing the efficacy CP treatment.

## Introduction

The emergence of severe acute respiratory syndrome coronavirus 2 (SARS-CoV-2) has led to a global pandemic with millions of infections reported across over 200 countries less than 6 months after the first case was reported [1–3]. A number of therapeutic agents are being evaluated, some of which are already in clinical use despite varying levels of evidence to support their efficacy [4]. One widely used agent is convalescent plasma (CP), the transfusion of plasma collected from individuals who have recovered from COVID-19 to currently infected patients.

In this context, CP is given with the hope of providing some degree of passive humoral immunity to the recipient via the transfer of antibodies directed against SARS-CoV-2 [5]. This approach has been used for centuries to treat infections, and more recent experiences with CP for other emerging viral infections suggest that CP may be an effective therapy for SARS-CoV-2 [6,7].

The evidence describing the efficacy of CP for patients with COVID-19 remains unclear. Early clinical reports from China on the use of CP for COVID-19 were encouraging [8–11], while another report suggests limited efficacy when used late in the course of disease [12]. Multiple randomized clinical trials are ongoing [13]. A report of early safety data from 20,000 patients given CP through a large expanded access protocol is reassuring [14].

This study describes the clinical outcomes of a cohort of hospitalized patients with severe COVID-19 who received CP. Notably, the study is one of the first to analyze the clinical outcomes of a large group of patients who received CP in comparison to a closely matched group receiving standard of care treatment.

## Methods

### Study setting and data collection

For this study we utilized data from adult patients admitted to three hospitals within the Lifespan health system, Rhode Island Hospital and The Miriam Hospital, both in Providence, Rhode Island, USA, and Newport Hospital, in Newport, Rhode Island, USA. This matched cohort study was an electronic chart review approved by the Institutional Review Board of RIH. All data were extracted from the electronic health record.

### CP characteristics and use

All patients who received CP at our institution did so through the expanded access protocol [15]. Due to limitations in locally available serologic testing at the time, CP was given to patients prior to knowing the SARS-CoV-2 antibody content. Instead, SARS-CoV-2 antibody content was assessed retrospectively on thawed segments (if available) using the Abbott Architect SARS-CoV-2 IgG assay (Abbott, Abbott Park, IL). All patients were prescribed 2 units of plasma and patients were included even if they only received one of the units.

In addition to the broad eligibility requirements set out in the CP expanded access protocol [16], patients were eligible to receive CP treatment if they also fulfilled the following local inclusion criteria: 1) symptom onset ≤ 10 days prior, 2) requiring supplemental oxygen (but not invasive ventilation), 3) no evidence of current hypercoagulability (D-dimer > 1000 µg/L, clinical signs of thrombosis).

### Control group patient selection

All adult patients with a positive molecular test for COVID-19 admitted to the hospital prior to May 31, 2020 who did not receive CP were reviewed for potential inclusion in the control group. To capture a similar case mix to those patients eligible for CP (see above), additional inclusion criteria for the control group included the following: 1) symptom onset ≤ 10 days prior to admission, 2) hospital admission ≥ 48 hours, 3) required supplemental oxygen (but not invasive ventilation) within 48 hours of hospitalization, 4) D-dimer obtained within 48 hours of hospitalization and < 1000 µg/L.

### CP group patient selection

All patients who received CP prior to May 31, 2020 were considered for potential inclusion in the CP group. Of note, our local practice regarding CP use (including implementation of our local inclusion criteria and increasing the dose of CP from 1 to 2 units) changed quickly after we started using this treatment option. The initial 10 patients given CP before this change were not included in this analysis. Several other patients who received CP were also excluded from the analysis after applying the control group inclusion criteria to the CP group. The decision to exclude these CP recipients was an effort to preserve uniformity within the CP group and between the CP and control groups and was made prior to any further data analysis.

### Study outcomes

The primary outcome of this study was the impact of CP treatment on all cause in-hospital mortality; the secondary outcome was the impact of CP treatment on the time to hospital discharge. All outcomes were censored at day 28.

### Statistical analysis

We compared patient’s characteristics between the two groups using Pearson’s Chi-square test for categorical variables, and Student’s t-test or Mann-Whitney-Wilcoxon test for continuous variable as means (with standard deviation, SD) or medians (with interquartile range, IQR), respectively. All analyses were performed using Stata v15.1 (Stata Corporation, College Station, TX). The statistically significant level was set at 0.05.

To evaluate the impact of CP treatment on all cause in-hospital mortality, we utilized both univariate and multivariate Cox proportional-hazards models. To evaluate the impact of CP treatment on time to hospital discharge, we used a stratified log-rank test and calculated the Mantel-Cox rate ratios. We also performed a subgroup analysis of in-hospital mortality and time to hospital discharge based on the antibody content of the CP units that each patient received.

## Results

### Study population and CP use

Of the 82 consecutive patients who received CP during the study period, 64 were included in the analysis. Excluded patients either received CP prior to the implementation of our local inclusion criteria or (in retrospect) did not meet the structured inclusion criteria for the matched control group [Figure 1]. Three patients included in the CP group received only 1 unit of CP (one withdrew due to clinical improvement and hospital discharge, two had transfusion-related acute lung injury (TRALI) reactions associated with the first unit). The remainder of the patients received 2 units of CP. The control group included 177 patients who did not receive CP [Table 1].

**Table 1.**
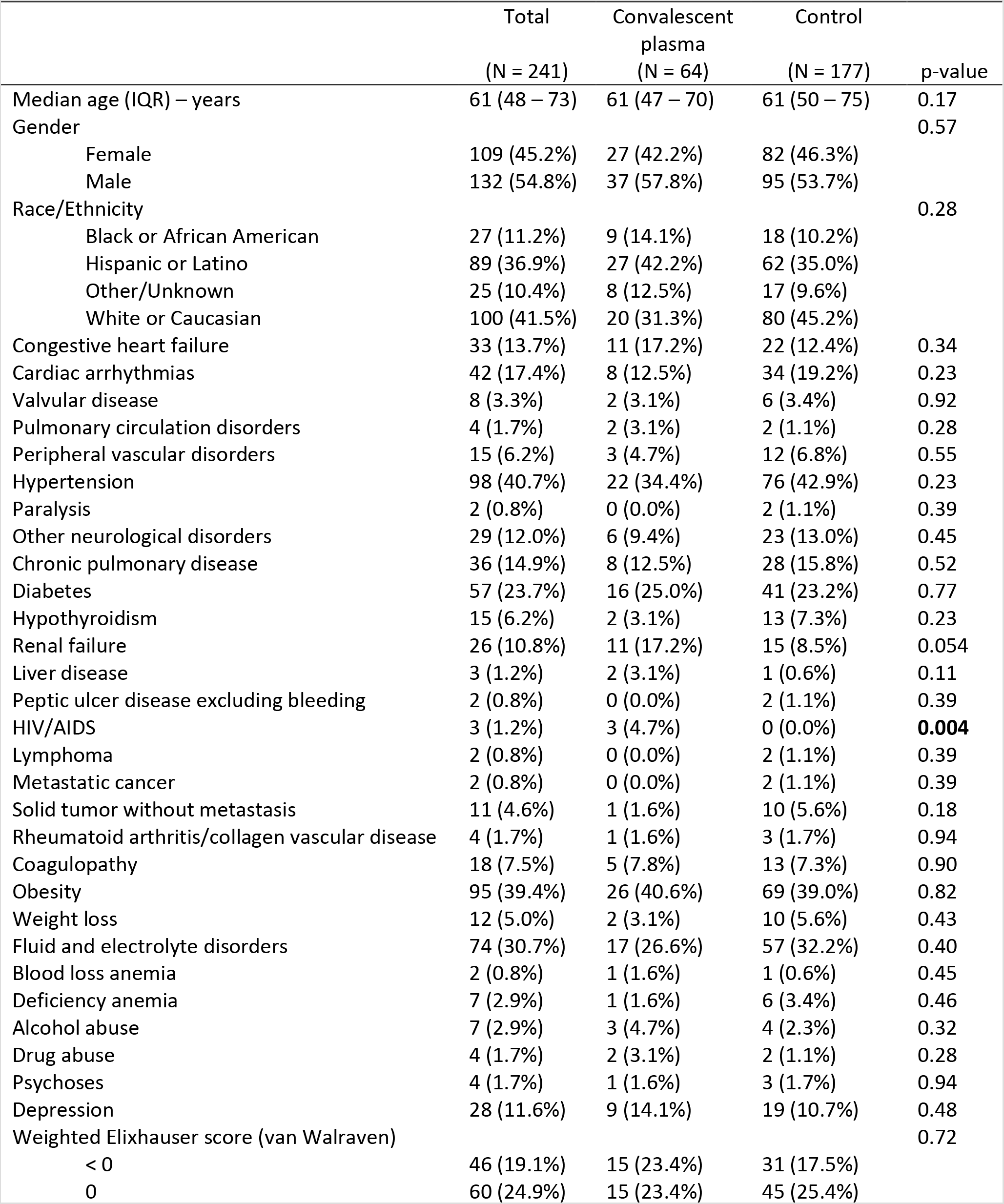
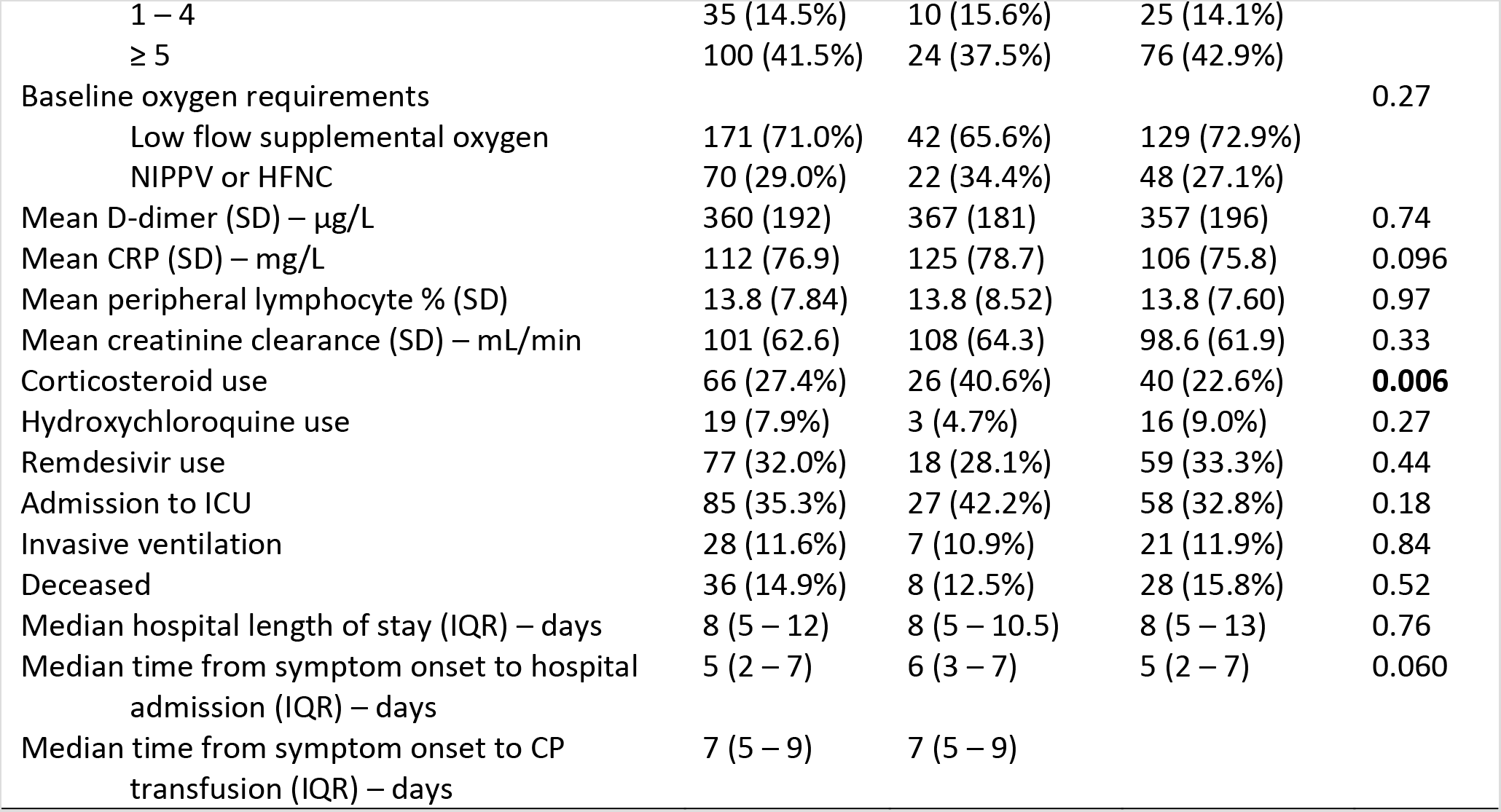
Patient characteristics.

|  | Total<br>(N = 241) | Convalescent<br>plasma<br>(N = 64) | Control<br>(N = 177) | p-value |
| --- | --- | --- | --- | --- |
| Median age (IQR) – years | 61 (48 – 73) | 61 (47 – 70) | 61 (50 – 75) | 0.17 |
| Gender |  |  |  | 0.57 |
| Female | 109 (45.2%) | 27 (42.2%) | 82 (46.3%) |  |
| Male | 132 (54.8%) | 37 (57.8%) | 95 (53.7%) |  |
| Race/Ethnicity |  |  |  | 0.28 |
| Black or African American | 27 (11.2%) | 9 (14.1%) | 18 (10.2%) |  |
| Hispanic or Latino | 89 (36.9%) | 27 (42.2%) | 62 (35.0%) |  |
| Other/Unknown | 25 (10.4%) | 8 (12.5%) | 17 (9.6%) |  |
| White or Caucasian | 100 (41.5%) | 20 (31.3%) | 80 (45.2%) |  |
| Congestive heart failure | 33 (13.7%) | 11 (17.2%) | 22 (12.4%) | 0.34 |
| Cardiac arrhythmias | 42 (17.4%) | 8 (12.5%) | 34 (19.2%) | 0.23 |
| Valvular disease | 8 (3.3%) | 2 (3.1%) | 6 (3.4%) | 0.92 |
| Pulmonary circulation disorders | 4 (1.7%) | 2 (3.1%) | 2 (1.1%) | 0.28 |
| Peripheral vascular disorders | 15 (6.2%) | 3 (4.7%) | 12 (6.8%) | 0.55 |
| Hypertension | 98 (40.7%) | 22 (34.4%) | 76 (42.9%) | 0.23 |
| Paralysis | 2 (0.8%) | 0 (0.0%) | 2 (1.1%) | 0.39 |
| Other neurological disorders | 29 (12.0%) | 6 (9.4%) | 23 (13.0%) | 0.45 |
| Chronic pulmonary disease | 36 (14.9%) | 8 (12.5%) | 28 (15.8%) | 0.52 |
| Diabetes | 57 (23.7%) | 16 (25.0%) | 41 (23.2%) | 0.77 |
| Hypothyroidism | 15 (6.2%) | 2 (3.1%) | 13 (7.3%) | 0.23 |
| Renal failure | 26 (10.8%) | 11 (17.2%) | 15 (8.5%) | 0.054 |
| Liver disease | 3 (1.2%) | 2 (3.1%) | 1 (0.6%) | 0.11 |
| Peptic ulcer disease excluding bleeding | 2 (0.8%) | 0 (0.0%) | 2 (1.1%) | 0.39 |
| HIV/AIDS | 3 (1.2%) | 3 (4.7%) | 0 (0.0%) | **0.004** |
| Lymphoma | 2 (0.8%) | 0 (0.0%) | 2 (1.1%) | 0.39 |
| Metastatic cancer | 2 (0.8%) | 0 (0.0%) | 2 (1.1%) | 0.39 |
| Solid tumor without metastasis | 11 (4.6%) | 1 (1.6%) | 10 (5.6%) | 0.18 |
| Rheumatoid arthritis/collagen vascular disease | 4 (1.7%) | 1 (1.6%) | 3 (1.7%) | 0.94 |
| Coagulopathy | 18 (7.5%) | 5 (7.8%) | 13 (7.3%) | 0.90 |
| Obesity | 95 (39.4%) | 26 (40.6%) | 69 (39.0%) | 0.82 |
| Weight loss | 12 (5.0%) | 2 (3.1%) | 10 (5.6%) | 0.43 |
| Fluid and electrolyte disorders | 74 (30.7%) | 17 (26.6%) | 57 (32.2%) | 0.40 |
| Blood loss anemia | 2 (0.8%) | 1 (1.6%) | 1 (0.6%) | 0.45 |
| Deficiency anemia | 7 (2.9%) | 1 (1.6%) | 6 (3.4%) | 0.46 |
| Alcohol abuse | 7 (2.9%) | 3 (4.7%) | 4 (2.3%) | 0.32 |
| Drug abuse | 4 (1.7%) | 2 (3.1%) | 2 (1.1%) | 0.28 |
| Psychoses | 4 (1.7%) | 1 (1.6%) | 3 (1.7%) | 0.94 |
| Depression | 28 (11.6%) | 9 (14.1%) | 19 (10.7%) | 0.48 |
| Weighted Elixhauser score (van Walraven) |  |  |  | 0.72 |
| < 0 | 46 (19.1%) | 15 (23.4%) | 31 (17.5%) |  |
| 0 | 60 (24.9%) | 15 (23.4%) | 45 (25.4%) |  |
| 1 – 4 | 35 (14.5%) | 10 (15.6%) | 25 (14.1%) |  |
| ≥ 5 | 100 (41.5%) | 24 (37.5%) | 76 (42.9%) |  |
| Baseline oxygen requirements |  |  |  | 0.27 |
| Low flow supplemental oxygen | 171 (71.0%) | 42 (65.6%) | 129 (72.9%) |  |
| NIPPV or HFNC | 70 (29.0%) | 22 (34.4%) | 48 (27.1%) |  |
| Mean D-dimer (SD) – µg/L | 360 (192) | 367 (181) | 357 (196) | 0.74 |
| Mean CRP (SD) – mg/L | 112 (76.9) | 125 (78.7) | 106 (75.8) | 0.096 |
| Mean peripheral lymphocyte % (SD) | 13.8 (7.84) | 13.8 (8.52) | 13.8 (7.60) | 0.97 |
| Mean creatinine clearance (SD) – mL/min | 101 (62.6) | 108 (64.3) | 98.6 (61.9) | 0.33 |
| Corticosteroid use | 66 (27.4%) | 26 (40.6%) | 40 (22.6%) | **0.006** |
| Hydroxychloroquine use | 19 (7.9%) | 3 (4.7%) | 16 (9.0%) | 0.27 |
| Remdesivir use | 77 (32.0%) | 18 (28.1%) | 59 (33.3%) | 0.44 |
| Admission to ICU | 85 (35.3%) | 27 (42.2%) | 58 (32.8%) | 0.18 |
| Invasive ventilation | 28 (11.6%) | 7 (10.9%) | 21 (11.9%) | 0.84 |
| Deceased | 36 (14.9%) | 8 (12.5%) | 28 (15.8%) | 0.52 |
| Median hospital length of stay (IQR) – days | 8 (5 – 12) | 8 (5 – 10.5) | 8 (5 – 13) | 0.76 |
| Median time from symptom onset to hospital admission (IQR) – days | 5 (2 – 7) | 6 (3 – 7) | 5 (2 – 7) | 0.060 |
| Median time from symptom onset to CP transfusion (IQR) – days | 7 (5 – 9) | 7 (5 – 9) |  |  |

**Figure 1.**
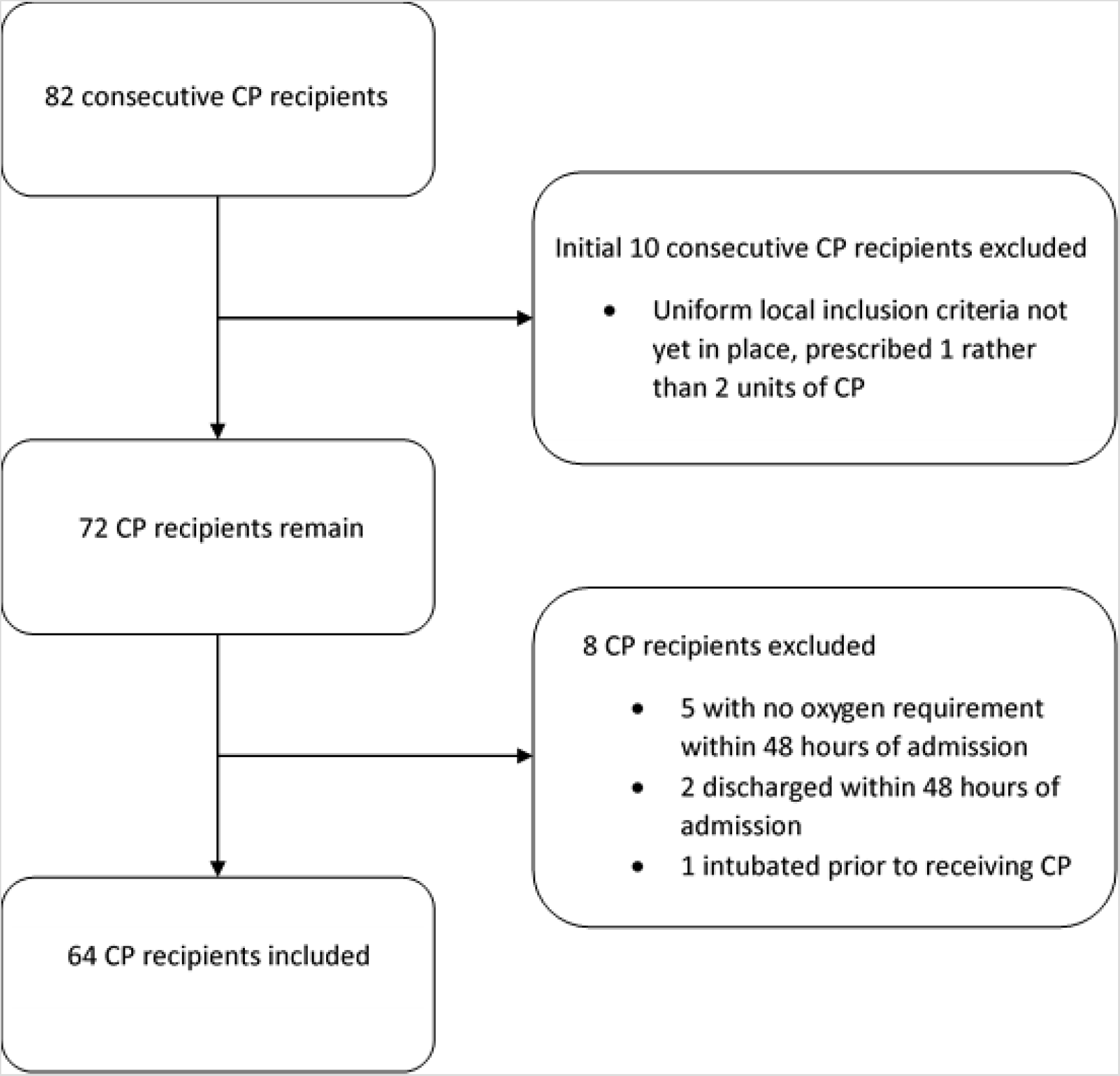
Flowchart for study inclusion of patients receiving convalescent plasma (CP).

Patients received CP at a median of 7 days after symptom onset. SARS-CoV-2 antibody testing was retrospectively performed on 97 (89.0%) of the 109 CP units which were transfused, 13 (13%) of these units had an antibody index (AI) below the cutoff for a positive result (AI < 1.4) [Figure 2]. All patients in the CP group except 3 received at least 1 unit of CP with an AI ≥ 1.4 and 18 patients received 2 units of CP both with AI ≥ 5 [Table 2].

**Table 2.** Convalescent plasma characteristics.

|  |  |
| --- | --- |
| Units of CP transfused | 109 |
| Units of CP with AI results available | 97 |
| AI < 1.4 | 13 (13%) |
| 1.4 ≤ AI < 5 | 27 (28%) |
| AI ≥ 5 | 57 (59%) |
| Patients transfused CP | 64 |
| No units with AI ≥ 1.4 | 3 (4.7%) |
| At least 1 unit with AI ≥ 1.4<sup>a</sup> | 32 (50%) |
| Two units both with AI ≥ 5.0 | 18 (28%) |
| Missing data | 11 (17%) |

Abbreviations: CP, convalescent plasma; AI, SARS-CoV-2 IgG antibody index

**Figure 2.**
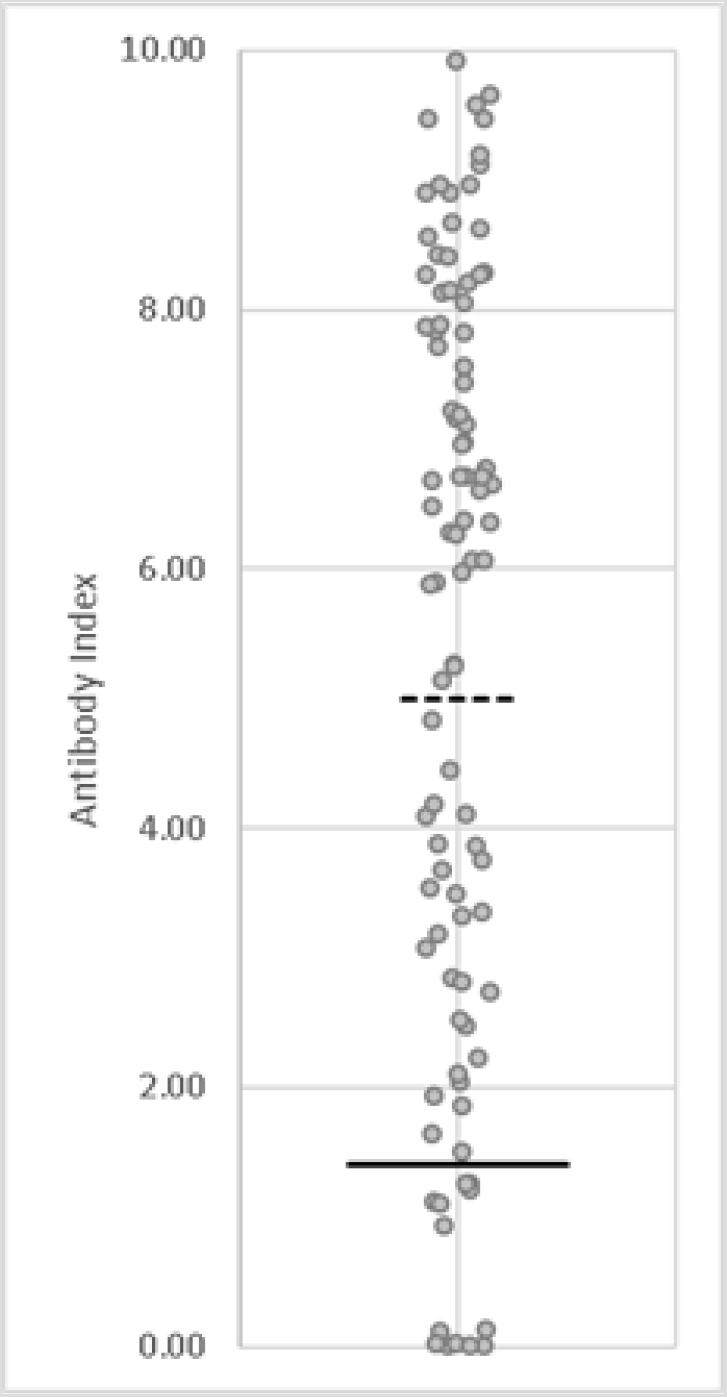
Distribution of SARS-CoV-2 IgG antibody index (AI) values detected in the convalescent plasma units transfused in this study. The red bar indicates the cutoff for a positive assay result (AI = 1.4) and the black bar indicates the cutoff for an arbitrarily high positive result (AI = 5).

### Clinical presentation and course

The demographics and summative pre-existing comorbidities score of the patients in the CP group and control groups were generally similar, although there were 3 patients with HIV/AIDS in the CP group (all of whom were on antiretroviral therapy) and none in the control group [Table 1].

Both the CP group and control group had a similar percentage of patients who were treated with remdesivir (28.1% v. 33.3%, p = 0.44), but the CP group had significantly more patients treated with corticosteroids (40.6% v. 22.6%, p = 0.006). Overall in-hospital mortality in this study was 14.9%, with 35.3% of patients admitted to the intensive care unit, and 11.6% requiring invasive ventilation. There was no significant difference between the groups in any of these categories (p = 0.52, p = 0.18, p = 0.84, respectively).

### Clinical outcomes and adverse events

The incidence of in-hospital mortality in the CP and control groups were not significantly different (12.5% v. 15.8%, p = 0.52) and a multivariate analysis also found no significant difference (adjusted hazard ratio [HR] 0.93, 95% confidence interval [CI] 0.39 – 2.20) [Table 3]. In a subgroup analysis examining only those patients who received 2 units of CP with AI ≥ 5, there was a lower risk for in-hospital mortality as compared to the control group, although this difference was also not significant (adjusted HR 0.39, 95% CI 0.05 – 3.08).

**Table 3.** Multivariate analysis of the impact of convalescent plasma treatment on all cause in-hospital mortality as compared to the control group.

| All cause in-hospital mortality | Unadjusted HR (95% CI) | Adjusted HR (95% CI) |  |
| --- | --- | --- | --- |
|  |  | Model 1<sup>a</sup> | Model 2<sup>b</sup> |
| Overall (n = 64) | 0.73 (0.32 – 1.69) | 0.91 (0.39 – 2.15) | 0.93 (0.39 – 2.20) |
| AI ≥ 1.4<sup>c</sup> (n = 32) | 1.08 (0.41 – 2.80) | 1.09 (0.41 – 2.86) | 1.17 (0.43 – 3.19) |
| AI ≥ 5<sup>d</sup> (n = 18) | 0.35 (0.05 – 2.62) | 0.38 (0.05 – 2.98) | 0.39 (0.05 – 3.08) |

Abbreviations: HR, hazard ratio; CI, confidence interval; AI, SARS-CoV-2 IgG antibody index
<sup>a</sup>Adjusted for age, gender, race, and baseline oxygen requirements.
<sup>b</sup>Adjusted for age, gender, race, baseline oxygen requirements, remdesivir use, and corticosteroid use.
<sup>c</sup>At least 1 unit with AI ≥ 1.4, but not 2 units both with AI ≥ 5
<sup>d</sup>Two units both with AI ≥ 5

The median length of stay in both the CP and control groups was 8 days. There was no significant difference in the overall rate of hospital discharge between the two groups using a stratified log-rank test (rate ratio [RR] 1.28, 95% CI 0.91 – 1.81) or in a subgroup analysis examining only those patients who received 2 units of CP with AI ≥ 5 as compared to the control group (RR 1.63, 95% CI 0.92 – 2.88) [Table 4].

**Table 4.** Stratified log-rank analysis of the impact of convalescent plasma treatment on time to hospital discharge as compared to the control group.

| | All CP recipients (n = 64)<br>RR (95% CI) | AI $\geq 1.4^a$ (n = 32)<br>RR (95% CI) | AI $\geq 5^b$ (n = 18)<br>RR (95% CI) |
| --- | --- | --- | --- |
| Overall | 1.28 (0.91 – 1.81) | 1.14 (0.72 – 1.83) | 1.63 (0.92 – 2.88) |
| Stratification subgroups |  |  |  |
| Sex |  |  |  |
| Female | 1.28 (0.75 – 2.19) | 1.31 (0.66 – 2.60) | 1.21 (0.40 – 3.68) |
| Male | 1.27 (0.80 – 2.00) | 1.00 (0.52 – 1.91) | 1.85 (0.94 – 3.64) |
| Age group (years) |  |  |  |
| 18 – 49 | 0.90 (0.48 – 1.70) | 1.77 (0.70 – 4.48) | 0.81 (0.29 – 2.29) |
| 50 – 64 | 0.82 (0.43 – 1.55) | 0.80 (0.37 – 1.75) | 1.57 (0.25 – 9.93) |
| $\geq 65$ | **1.86 (1.03 – 3.36)** | 1.28 (0.58 – 2.85) | **2.70 (1.16 – 6.28)** |
| Race/ethnicity |  |  |  |
| Black or African American | 1.49 (0.56 – 3.93) | 1.19 (0.34 – 4.14) | 3.00 (0.67 – 13.4) |
| Hispanic or Latino | 0.88 (0.51 – 1.54) | 1.09 (0.48 – 2.51) | 0.95 (0.44 – 2.05) |
| Other/Unknown | 1.38 (0.54 – 3.51) | 1.75 (0.44 – 6.91) | 1.20 (0.14 – 10.5) |
| White or Caucasian | 1.51 (0.82 – 2.76) | 1.05 (0.52 – 2.14) | **6.67 (1.39 – 32.1)** |
| Baseline oxygen requirements |  |  |  |
| Low flow supplemental oxygen | 1.34 (0.89 – 2.03) | 1.15 (0.68 – 1.94) | 2.03 (0.90 – 4.56) |
| NIPPV or HFNC | 1.52 (0.78 – 2.96) | 1.00 (0.33 – 3.02) | 2.36 (0.97 – 5.74) |
| Days from symptom onset to admission |  |  |  |
| $\leq 5$ | 1.31 (0.79 – 2.16) | 1.03 (0.52 – 2.04) | 1.82 (0.66 – 5.06) |
| $> 5$ | 1.16 (0.71 – 1.89) | 1.21 (0.63 – 2.35) | 1.30 (0.64 – 2.65) |
| Remdesivir use |  |  |  |
| No | 1.20 (0.79 – 1.82) | 1.04 (0.60 – 1.78) | 1.66 (0.83 – 3.33) |
| Yes | 1.41 (0.75 – 2.66) | 1.68 (0.63 – 4.50) | 1.37 (0.50 – 3.77) |
| Corticosteroid use |  |  |  |
| No | 1.25 (0.81 – 1.93) | 0.97 (0.52 – 1.83) | 1.94 (0.94 – 4.01) |
| Yes | 1.66 (0.90 – 3.05) | 1.74 (0.82 – 3.69) | 1.66 (0.63 – 4.37) |

Abbreviations: RR, rate ratio; CI, confidence interval; AI, SARS-CoV-2 IgG antibody index; NIPPV, non-invasive positive pressure ventilation; HFNC, high-flow nasal cannula
<sup>a</sup>At least 1 unit with AI $\geq 1.4$ , but not 2 units both with AI $\geq 5$
<sup>b</sup>Two units both with AI $\geq 5$

Patients 65-years-old or greater who received CP had an increased rate of hospital discharge as compared to those who did not (RR 1.86, 95% CI 1.03 – 3.36); this increased rate of hospital discharge was even more pronounced in a subgroup analysis of elderly patients who received 2 units of CP with AI ≥ 5 (RR 2.70, 95% CI 1.16 – 6.28). There was also a significant difference in the rate of hospital discharge for White or Caucasian patients who received 2 units of CP with AI ≥ 5.

Two patients who received CP were judged to have a TRALI reaction. The first, a previously healthy 37-year-old male, had severe chest pain 45 minutes after starting the CP transfusion with associated tachypnea and worsening hypoxemia, the transfusion was stopped and symptoms resolved several hours later. The second, a 36-year-old male who had recently undergone kidney transplantation for end-stage renal disease, also had acute worsening of tachypnea and hypoxemia associated with chest pain approximately 30 minutes after starting the CP transfusion, the transfusion was stopped and symptoms resolved within 24 hours. Both patients remained afebrile and without elevated BNP during these episodes. Additionally, a 64-year-old female with end-stage renal disease on hemodialysis who received CP was judged to have transfusion-associated circulatory overload (TACO) with acutely worsening hypoxemia approximately 3 hours after transfusion of 2 units of CP. There were no other documented adverse events associated with CP use for the patients included in this study.

## Discussion

This matched cohort study examined the use of CP in hospitalized patients with severe COVID-19 and found no significant difference in overall in-hospital mortality or time to hospital discharge as compared to a control group who did not receive CP. The CP group and control groups in this study were drawn from a well-defined population, were similar in baseline characteristics and severity of illness, and were followed long enough to allow for adequate analysis of clinical outcomes. We found a signal for the potential efficacy of CP in elderly patients, a signal for a larger potential effect of CP with a higher quantity of measured SARS-CoV-2 antibody, and a greater than expected frequency of transfusion reactions.

Importantly, the stratified analysis examining only patients 65-years-old or greater who received CP showed a significantly increased rate of hospital discharge as compared to the control group. An effect specific to this age group is not entirely surprising given the increase in morbidity among the elderly with COVID-19, the waning of humoral immunity with age, and the importance of the humoral compartment of the overall immune response in combating this infection [17–19]. If this increased efficacy of CP in the elderly is re-demonstrated in other settings, this may impact the design of clinical trials for SARS-CoV-2 monoclonal antibodies or aid in the triage of limited CP resources.

A subgroup analysis examining only those patients who received 2 units of CP with AI ≥ 5 showed an even larger increase in the rate of hospital discharge among the elderly as compared to the control group. There was also a statistically significant increase in the rate of hospital discharge among White/Caucasian patients of all ages, although this may have been primarily driven by the relative increased age of the White/Caucasian patients in our study as compared patients of other races or ethnicities (data not shown). The semi-quantitative description of the amount of SARS-CoV-2 anti-nucleocapsid IgG provided by the assay’s antibody index (AI) that was used in this study has been shown to have similar positivity rate as a recombinant neutralizing assay, although the correlation between neutralizing titers and AI was demonstrated to be poor [20]. In a more recent study using a SARS-CoV-2 neutralizing assay, a different SARS-CoV-2 anti-spike IgG assay was very well correlated with the neutralizing titer [21]. Although we cannot draw any direct conclusions about the neutralizing titers of the CP used in our study, the increase in the rate of hospital discharge across multiple stratifications seen in the subgroup with AI ≥ 5 as compared to the entire CP group aligns well with the expected and recently demonstrated dose-dependent effect of CP [22].

There was a greater than expected frequency of transfusion reactions in the CP group. The observed per unit reaction rate of 3/109 (2.8%) is consistent with the 2.5% reported by Li et al [23] and higher than that reported to our blood banks (< 1%, data not shown). We classified two cases as TRALI reactions. We are aware that CP is manufactured from never transfused male donors and never pregnant females or the CP is tested for anti-HLA antibodies. Hence, neither product (donor) was investigated for anti-HLA antibodies. However, the pathophysiology of Type 2 TRALI in hospitalized patients is not related to donor alloantibodies, but to other compounds with inflammatory evoking properties in the context of an activated pulmonary endothelium, which clearly pertains to COVID-19 patients [24]. Our elevated frequency of transfusion reactions does not align with data reported from the large safety study associated with the expanded access protocol (TRALI 0.10%, TACO 0.18%, severe allergic transfusion reaction 0.13%) [14]. This difference may highlight the difficulty of accurately detecting transfusion reactions in critically ill patients or may instead simply represent an overestimation of our local reaction rate due to the relatively low number of patients we have transfused thus far.

Studying the efficacy of CP for emerging infectious diseases in an adequately powered prospective randomized fashion has historically been difficult. Retrospective or non-randomized assessments of efficacy cannot provide the same quality of evidence, as highlighted by the contrasting data provided by a non-randomized cohort study of CP for severe pandemic influenza A (H1N1) 2009 which suggested a mortality benefit of adding CP to the local standard of care [25], and two more recent well powered randomized controlled trials (RCTs) examining the use of high-titer anti-influenza plasma as compared to placebo [26] or low-titer plasma [27] for the treatment of seasonal influenza infection, neither of which showed a significant difference in the primary outcome studied (clinical status at day 7). In light of these findings, a critical analysis of any non-randomized study assessing the efficacy of CP is imperative.

While our study does demonstrate a lower incidence of in-hospital mortality in the CP group as compared to the control group, this difference was not significant, and a multivariate analysis also showed no significant difference between the groups. Two other propensity-matched cohort studies suggest a mortality benefit of CP for severe COVID-19 [28,29], while two (perhaps underpowered) RCTs were unable to demonstrate any mortality benefit [23,30]. The timing of CP transfusion in relation to symptom onset, the antibody titer of the CP transfused, and the quantity of plasma transfused all varied widely among these studies, making it difficult to analyze their outcomes collectively, a topic explored in several recent meta-analyses [31–33]. The initial outcomes data from over 35,000 patients who received CP under the expanded access protocol reemphasizes the importance of these factors, with a high CP antibody titer and earlier transfusion corresponding to decreased mortality, as compared to a lower titer or later transfusion [22].

The findings of this study should be generalized with caution. There is the possibility of an unknown confounder biasing the composition of the CP group and the control group. Many of the patients in the control group were hospitalized either before CP was locally available or during the peak of COVID-19 hospitalizations when logistical constraints may have prevented them from being offered CP. To avoid these confounding effects, the control group was constructed to contain patients with similar characteristics as the CP group by applying the inclusion criteria for CP to all patients hospitalized with COVID-19 during the study period. The CP and control groups were generally well matched among all variables examined except corticosteroid use (significantly higher in the CP group), but both the multivariate mortality analysis and stratified rate of hospital discharge analysis accounted for this difference. The single center nature of the study allows for a comparison with a well-matched controlled group but also limits enrollment, and thus the study may have been underpowered to show a significant difference in the outcomes studied. We also did not specifically analyze an outcome examining time to clinical improvement and using hospital discharge as a surrogate for this measure may be misleading since some patients may have remained hospitalized for unrelated reasons.

In conclusion, this is one of the first analyses comparing CP to standard of care for the treatment of COVID-19. Though our study had several limitations, we found no overall difference in in-hospital mortality or in time to hospital discharge for those patients who received CP as compared to those who did not. A secondary analysis showed a significantly increased rate of hospital discharge for CP given to patients 65-years-old or greater. While manufactured hyperimmune globulin or monoclonal antibodies may eventually supplant CP in the developed world, CP may remain a more cost-effective and feasible option in the developing world as it can be manufactured locally. Moving forward, it will be important to keep in mind a relevant transfusion medicine principle: give the right blood (plasma containing an adequate concentration of antibody) to the right patient (perhaps older adults or those with immunocompromising conditions) at the right time (early in the course of disease). We anxiously await the results of several ongoing randomized trials of CP taking place across the globe to help gain further insight into this therapy.

## Data Availability

The data underlying this article are available in the article.

## Abbreviations

AI: antibody index
COVID-19: coronavirus disease 2019
CI: confidence interval
CP: convalescent plasma
HFNC: high-flow nasal cannula
HR: hazard ratio
ICU: intensive care unit
IQR: interquartile range
NIPPV: non-invasive positive pressure ventilation
RR: rate ratio
SARS-CoV-2: severe acute respiratory syndrome coronavirus 2
SD: standard deviation
TACO: transfusion-associated circulatory overload
TRALI: transfusion-related acute lung injury

